# Pitting the Gumbel and logistic growth models against one another to model COVID-19 spread

**DOI:** 10.1101/2020.05.24.20111633

**Authors:** Keunyoung Yoo, Mohammad Arashi, Andriette Bekker

**Author notes:** Corresponding author: Department of Statistics, Faculty of Natural and Agricultural Sciences, University of Pretoria, South Africa. E-mail addresses.

## Abstract

In this paper, we investigate briefly the appropriateness of the widely used logistic growth curve modeling with focus on COVID-19 spread, from a data-driven perspective. Specifically, we suggest the Gumbel growth model for behaviour of COVID-19 cases in European countries in addition to the United States of America (US), for better detecting the growth and prediction. We provide a suitable fit and predict the growth of cases for some selected countries as illustration. Our contribution will stimulate the correct growth spread modeling for this pandemic outbreak.

## Graphical abstract

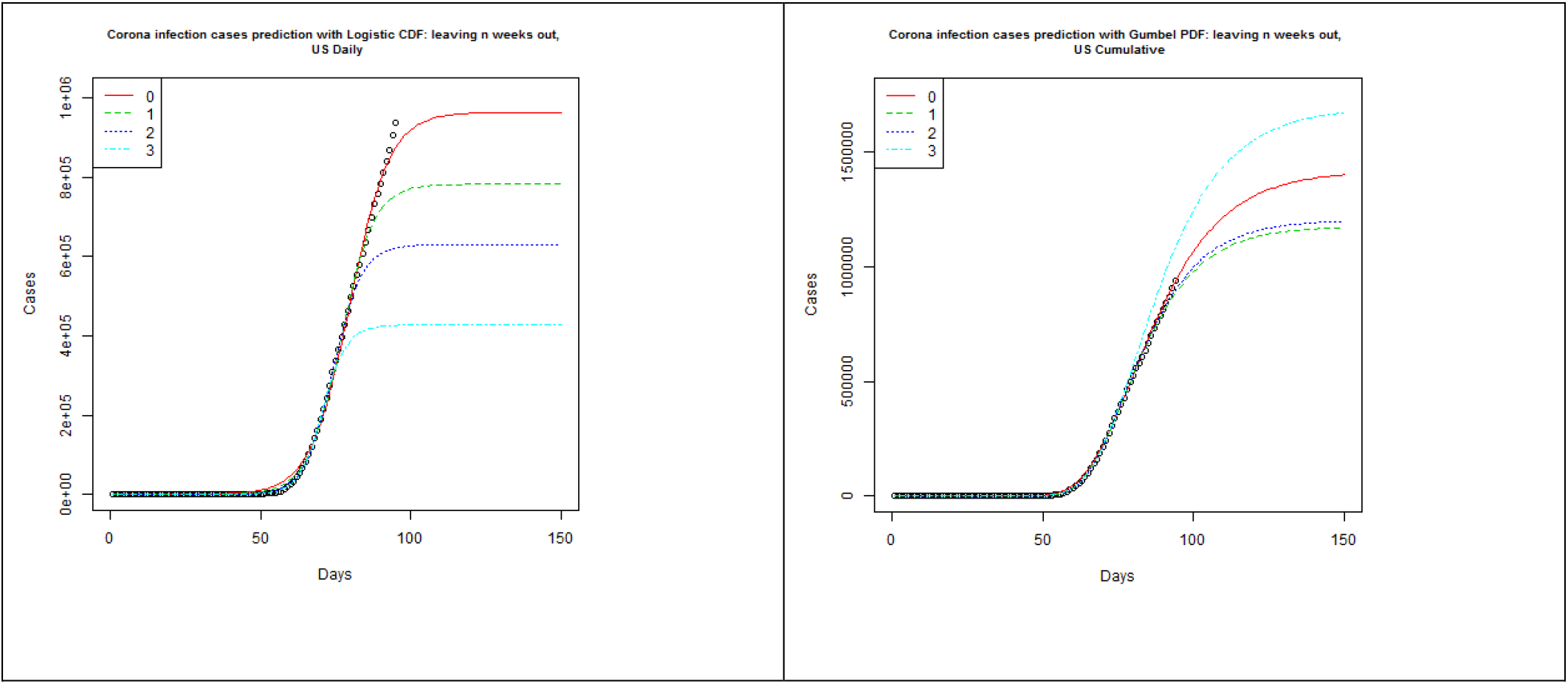

## 1. Introduction

Nowadays, the Coronavirus pandemic, known as COVID-19, caused by a novel pathogen named Severe Acute Respiratory Syndrome Coronavirus 2 (SARS-CoV2), has shown that in early stages of infection, symptoms of severe acute respiratory infection can occur and it is rapidly spreading across the globe. Since we have limited knowledge about COVID-19, epidemiological modeling is still under development and modeling the ecological growth based on the population demographic information is feasible for reporting. It is to support the shaping of decisions around different non-pharmaceutical interventions.

The logistic function/curve is commonly used for dynamic modeling in many branches of science including chemistry, physics, material science, forestry, disease progression, sociology, etc. But, the question is whether it is also suitable for COVID-19 spread modeling from the available data viewpoint. The principle of exponential growth can be applied to the transmission of COVID-19 (see [1] for a web based dashboard). It is known that the exponential model is adequate to describe for a short period and in general it will quickly deviate from actual numbers as time passes. The logistic growth curve was successful in modeling some epidemics (see [2], [3], [4], [5] and [6]). Our primary goal is to see whether the logistic function can suitably predict the spread. Some endeavours have been made to predict and forecast the future trajectory of the COVID-19 outbreak. We refer to [7], [8], [9], [10], [11], [12], [13], [14], [15], [16] and [17] to mention a few related studies.

In none of the above mentioned studies, the Gumbel function is applied for predicting the growth of COVID-19. Hence, in this contribution, a dynamic Gumbel model is used to track the coronavirus COVID-19 outbreak. We organize the rest of this work as follows. In the forthcoming section, we provide the source of data and software used for comparison and fitting purposes. Section 2 includes the analysis of logistic modeling, outlines the shortcomings, proposes the Gumbel model as the suitable candidate; followed by comparison with the Logistic model. Section 3 illustrates the potential of the Gumbel model with the analysis of the COVID-19 data for selected countries. We conclude our contribution in Section 4. The highlights of this paper can be summarised as follows:

- Exploring logistic curve modeling for COVID-19 data and illustrating the shortcomings.
- Proposing the modeling of COVID-19 spread with the Gumbel growth curve.
- Fitting of COVID-19 data from different countries to strongly support the Gumbel model choice.

### 1.1. Experimental data

There are a number of sources on the web that provide data on COVID-19 cases. One such site is “The Humanitarian Data Exchange” and one can find daily cumulative cases of COVID-19 per country. https://data.humdata.org/dataset/novel-coronavirus-2019-ncov-cases has a downloadable “time_series_covid19_confirmed_global.csv” starting from 2020-01-22, and for some countries, it even has the data broken down into different states or provinces. In order to perform the desired analysis, daily cases for each country had to be obtained, but some countries, such as the US and Australia, had the data broken down to state or provincial level. Since the focus of this research was per country, R open source software was used to sum along the unique values of Country, appropriately transforming the data for our analysis, then and non-linear regression was performed using the nls function.

## 2. Methodology and Results

### 2.1. Preliminary insight by using the logistic growth model

In this section, we conduct data analysis using the commonly used logistic curve modeling. A logistic function is a common sigmoid curve with the following functional form for the dynamic model of population at time *t*

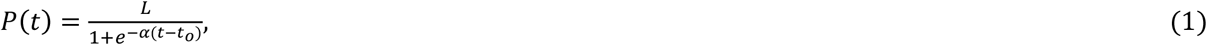

with initial condition *P(t_o_) = P_o_*, *L* is the carrying capacity, the maximum capacity of the environment here, *α* > 0. Here, Eq. (1) divided by *L* corresponds to the cumulative distribution function (CDF) of a logistic distribution at point *t*. The probability density function (PDF) is simply obtained by differentiating the latter with respect to *t*.

This is useful because the difference of two Gumbel-distributed random variables has a logistic distribution. The seemingly exponential growth of COVID-19 cases across the globe is typically the lower half of a logistic curve during the early stage.

#### 2.1.1. Results and discussion-Logistic model

The analysis is data-driven, and therefore, the focus of the paper is not from an epidemiological perspective. Nevertheless, the parameter estimates are relatable to the real world. *L* represents how many cases we expect to see in the end, *α* is how quickly the virus has spread/cleared and *t_o_* is where the peak increase in cases was observed. To illustrate the failing of the logistic model, the US data was the focus here.

Modeling the US cases, based on data until 28 March, the following results were obtained for regression. The model was highly significant with a p-value less than 0.0001 and this is shown in the plot as well, where the actual US data and the model are almost indistinguishable. This data suggests the total number of COVID-19 cases will be approximately between 226,000 and 265,000. The number of cases for the next 7 weeks was forecasted using these estimates. However, when data until 4 April is subsequently used, parameter *L*, which represents the final number of cases (477922) is far beyond what was predicted using data until the previous week (upper bound for the confidence limit of a was 265206). The slope parameter, *α*, decreased while the location parameter, *t_o_* increased. (See Fig 1 and Fig 2.).

**Fig 1.**
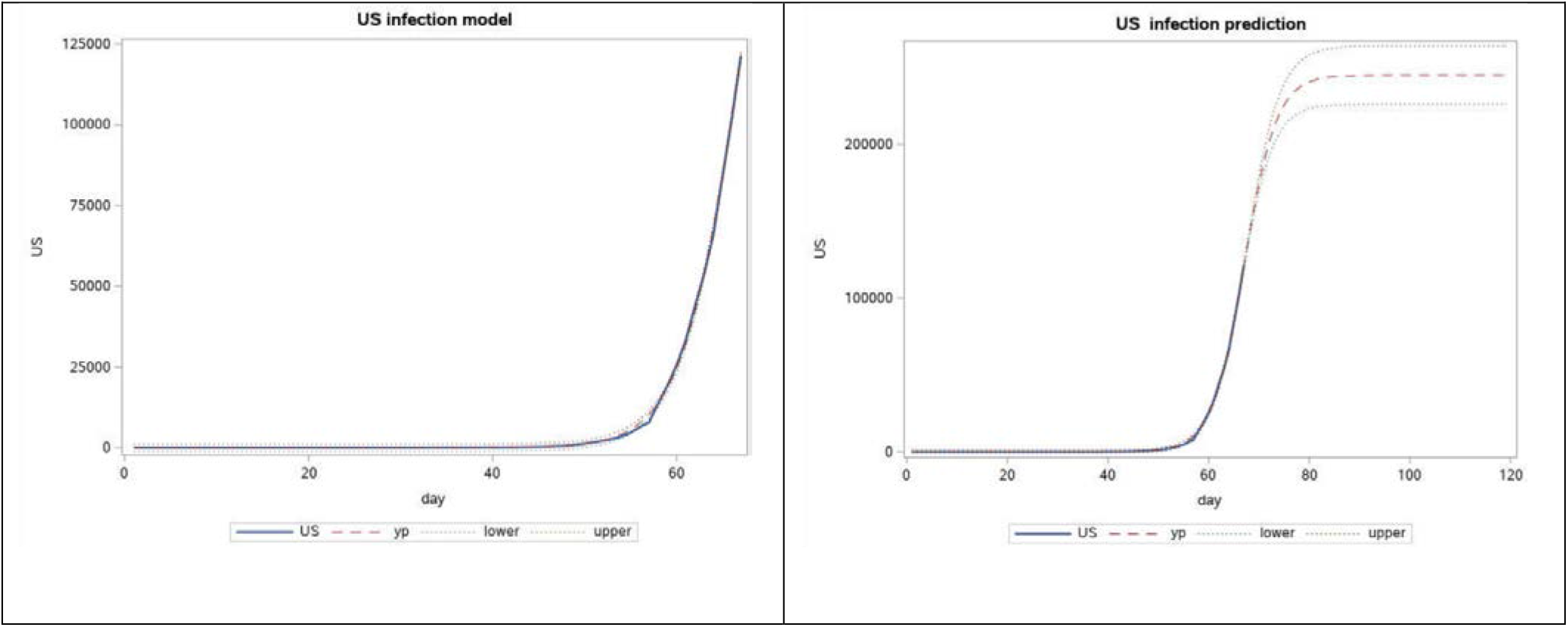
Observed US cases from 2020-01-22 to 03-28 and forecast for 7 weeks, using logistic function.

**Fig 2.**
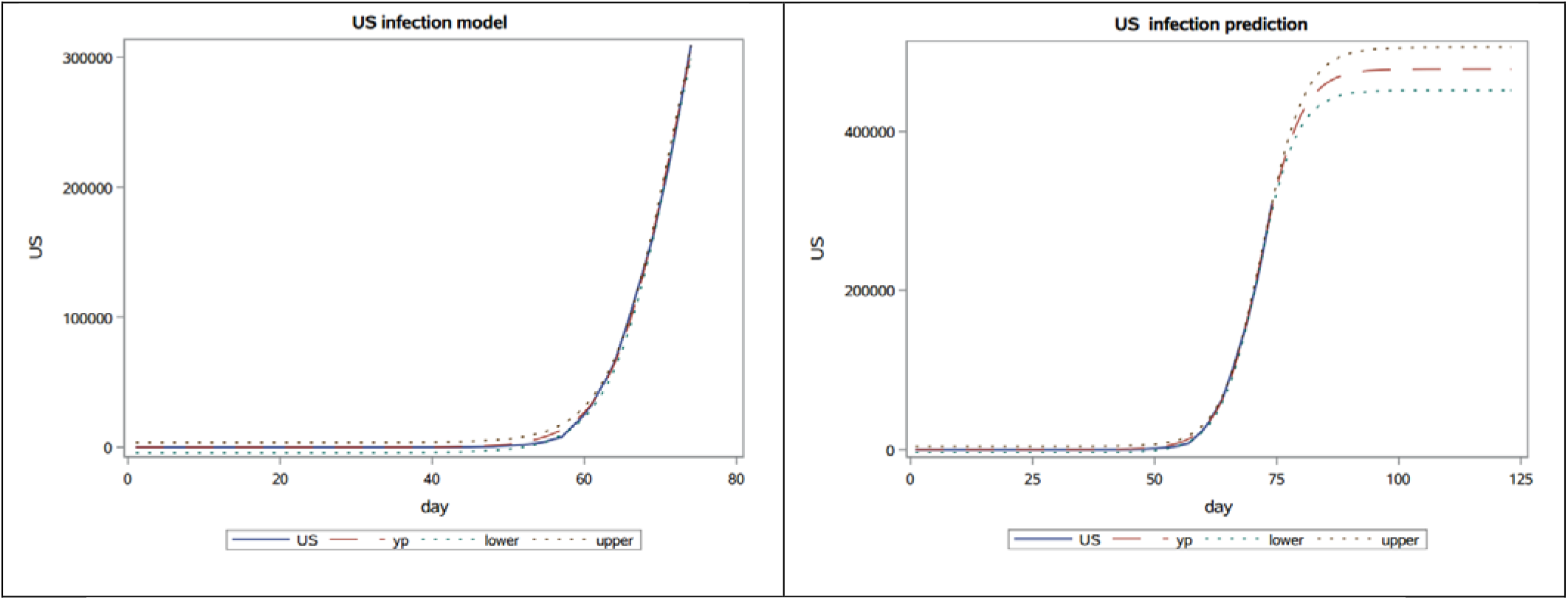
Observed US cases from 2020-01-22 to 04-04 and forecast for 7 weeks, using logistic function.

Using data until 2020-04-25, the new estimate for *L* once again exceeds what was predicted using previous data and the slope parameter, *α*, decreased while the location parameter, *t_o_* increased. (See Fig 3.)

**Fig 3.**
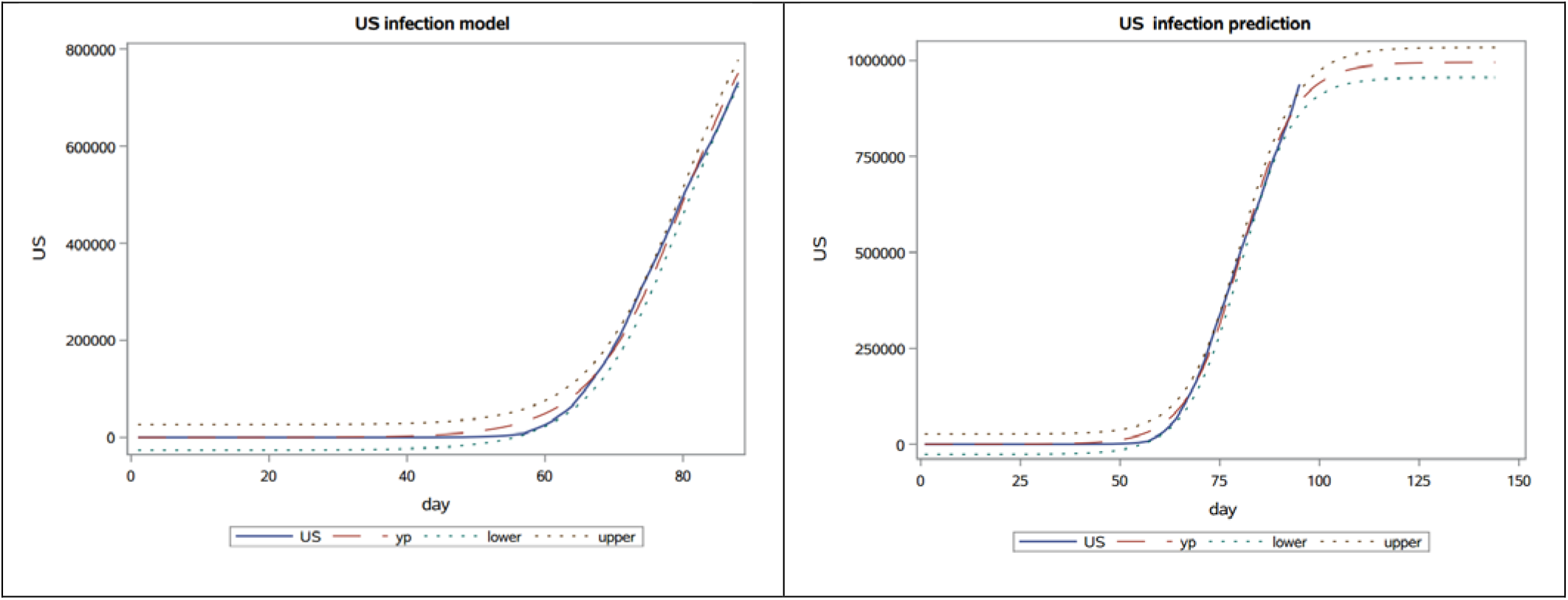
Observed US cases from 2020-01-22 to 04-25 and forecast for 7 weeks, using logistic function.

**Table 1.** US- logistic function based parameter estimates from 2020-01-22 to 03-28.

| Parameter | Estimate | Approx Standard Error | Approx 95% Confidence Limits |  |
| --- | --- | --- | --- | --- |
| $L$ | 245898 | 9653.8 | 226590 | 265206 |
| $\alpha$ | 0.3098 | 0.00483 | 0.3001 | 0.3195 |
| $t_o$ | 67.1124 | 0.2341 | 66.6442 | 67.5806 |

**Table 2.** US- logistic function based parameter estimates from 2020-01-22 to 04-04.

| Parameter | Estimate | Approx Standard Error | Approx 95% Confidence Limits |  |
| --- | --- | --- | --- | --- |
| $L$ | 477922 | 13319.9 | 451282 | 504562 |
| $\alpha$ | 0.2403 | 0.00405 | 0.2322 | 0.2484 |
| $t_o$ | 71.7055 | 0.2409 | 71.2236 | 72.1874 |

**Table 3.** US- logistic function based parameter estimates from 2020-01-22 to 04-25.

| Parameter | Estimate | Approx Standard Error | Approx 95% Confidence Limits |  |
| --- | --- | --- | --- | --- |
| $L$ | 995370 | 14689.3 | 966196 | 1024544 |
| $\alpha$ | 0.1459 | 0.00321 | 0.1395 | 0.1523 |
| $t_o$ | 80.3266 | 0.2772 | 79.7761 | 80.8772 |

Modelling the cumulative cases can be viewed as trying to model the forest as a whole, as opposed to looking at each tree. Even if a particular tree is twice as tall as most other trees in the forest, it will not make a big impact on the whole when all the heights are summed up. Therefore, to introduce more variability to the data, the next approach was to analyse the daily new cases instead, by taking the difference of the cumulative data. This way, the magnitude of daily cases will not be reduced as more data is acquired, and it will capture the effect of large spikes. In other words, we are zooming into the data to give more weight to the daily number of cases.

The parameter estimates below are based on the same data as the one above. The p-value for the model was still <0.0001, suggesting that its significance was not lost in the new approach. One observation was lost in the process of taking the difference, but by looking at the daily cases and trying to fit a PDF instead of a CDF, we can get a much detailed view of the situation, and it has increased the estimate for the total number of cases. See Fig 4; the view by focusing on daily case modelling using the first derivative of the logistic function.

**Fig 4.**
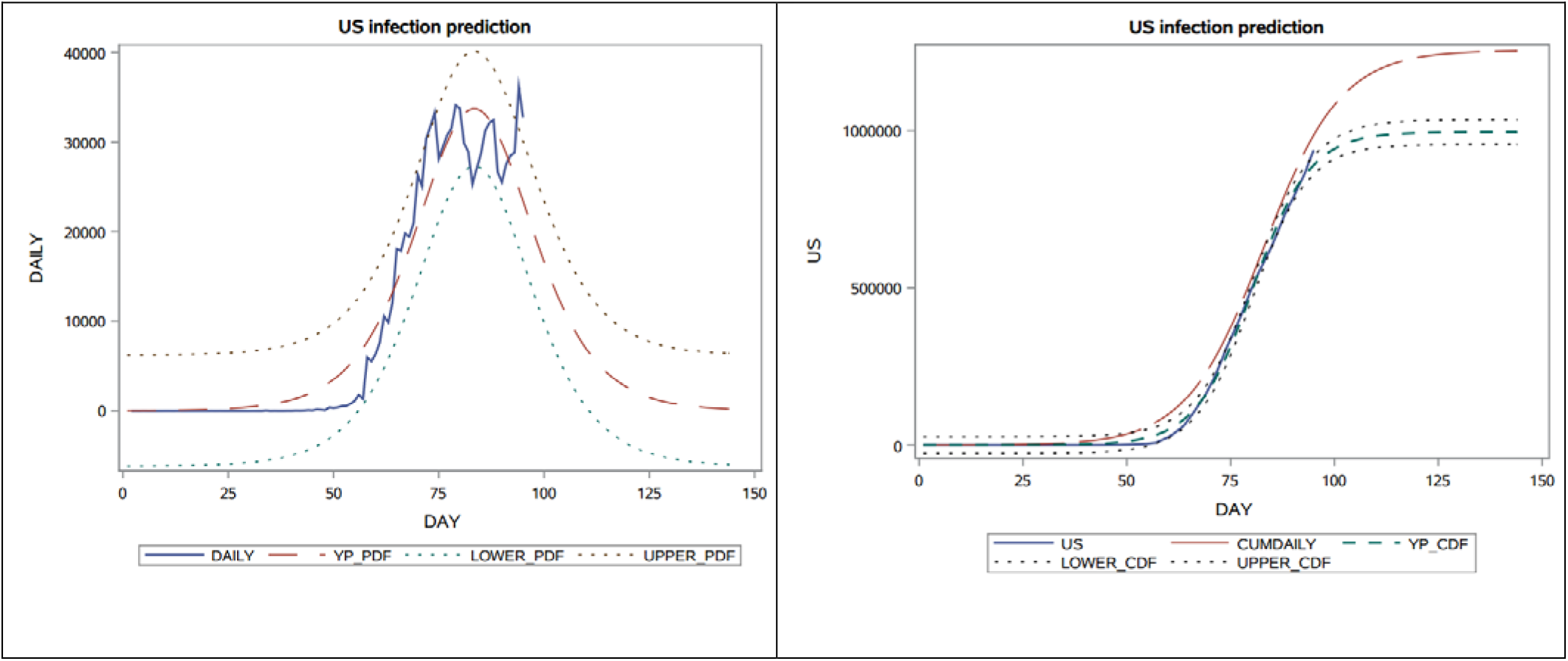
Observed daily US cases from 2020-01-22 to 04-25 and forecast for 7 weeks, using logistic function’s derivative.

**Table 4.** US- logistic function’s derivative based parameter estimates from 2020-01-22 to 04-25.

| Parameter | Estimate | Approx Standard Error | Approx 95% Confidence Limits |  |
| --- | --- | --- | --- | --- |
| $L$ | 1254903 | 47672.7 | 1160207 | 1349599 |
| $\alpha$ | 0.1076 | 0.00493 | 0.0978 | 0.1174 |
| $t_o$ | 83.4375 | 0.5791 | 82.2871 | 84.5879 |

#### 2.1.2. Shortcomings

In using a sigmoid function to model the data, an implicit assumption was made that it will take the same length of time for the spread of virus to “rise” as it will to “fall.” This comes from the fact that the Logistic function is symmetrical about the inflection point. The bar charts (see Fig. 5) show the daily new cases for Spain, Italy and the US. Just looking at the charts below is enough to question whether trying to fit a symmetrical shaped curve will provide a good fit or predictability. Hence the next step was to find a distribution whose CDF appears to have the general “S” shape which has the characteristics of a sigmoid function, yet possesses some skewness built into it such that when modelling the daily new cases, it fits the asymmetrical data well. After looking at numerous distributions that meet all criteria, the Gumbel distribution seemed to possess promising properties.

**Fig 5.**
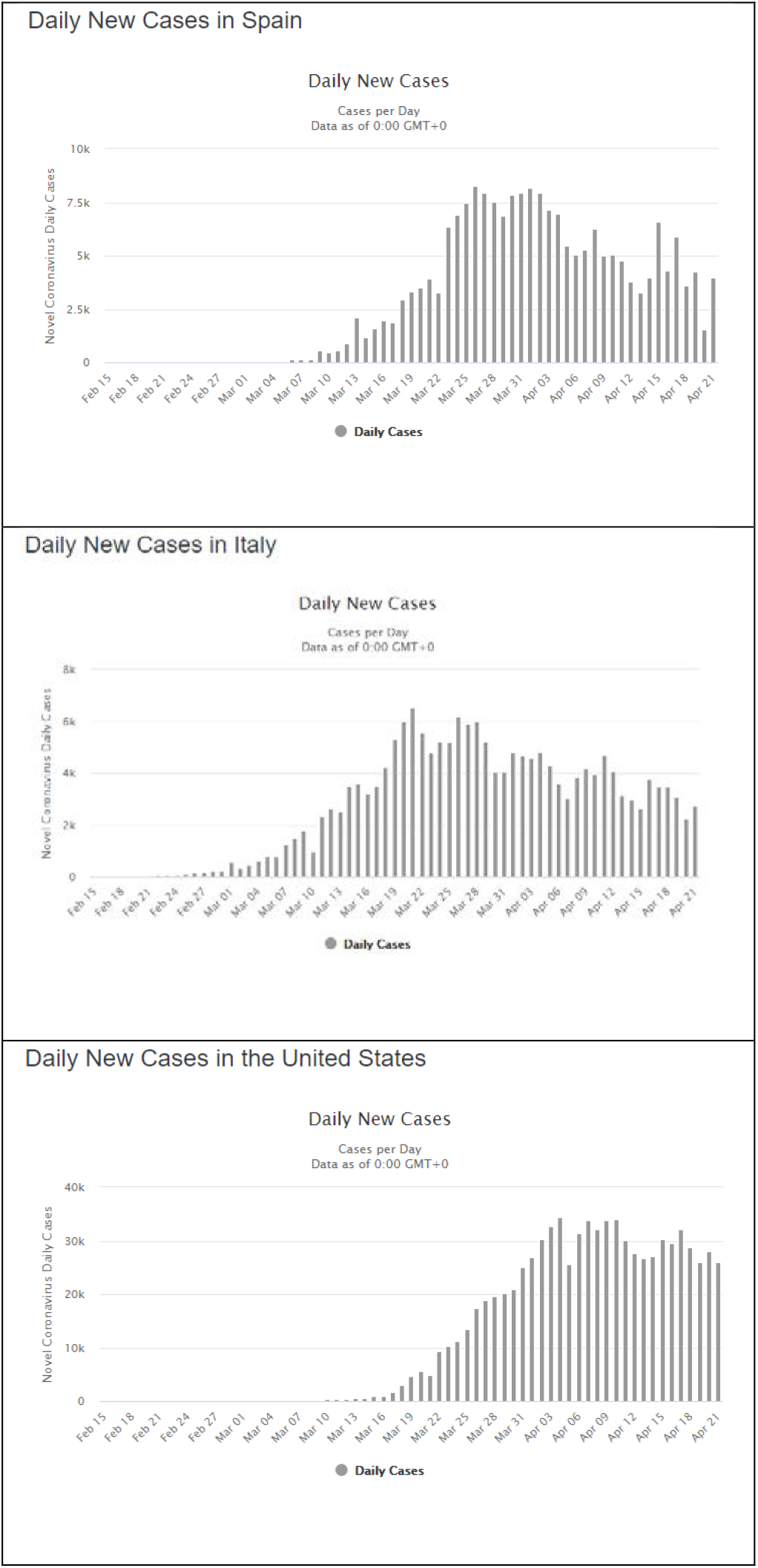
Bar chart of Daily new cases in Spain, Italy and the US (extracted from https://www.worldometers.info/coronavirus/country/us/)

Fig 6 suggests how easily our eyes can deceive us. The red lines (CDF and PDF) are the Logistic distribution and the blue lines (CDF and PDF) are the Gumbel distribution. (The dashed lines are the PDF and the solid lines are the CDF.) If we were to just view the CDFs in isolation, there is no way that a human will be able to tell whether the curve is symmetric or not. Even with the x and y axis drawn, merely shifting the Gumbel CDF to the left slightly will be enough to fool the viewer that the distribution is convincingly symmetric. On the other hand, detecting symmetry (or lack thereof) using a PDF is visually clear, and it does not require an expert to determine that while the dashed red curve (of the Logistic function) is symmetric, the dashed blue curve (of Gumbel) is not. Hence looking at the daily data and detecting this skewness was crucial in suggesting an alternative model.

**Fig 6.**
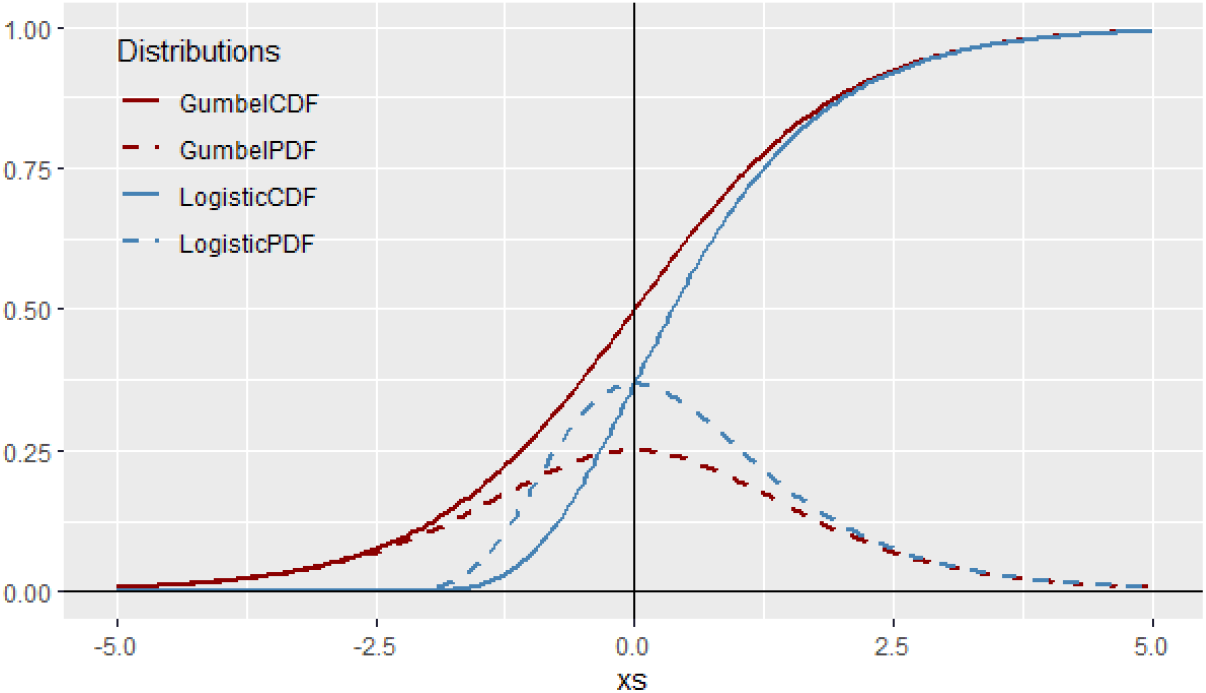
PDF and CDF of Logistic distribution and Gumbel distribution.

### 2.2. Gumbel growth modeling

The Gumbel distribution has been frequently used for practical probabilistic modeling. Gumbel ([18], [19], [20] and [21]) presents a model as an extension of the exponential distribution with the feature that it can be used to fit extreme datasets. A Gumbel dynamic model of population at time *t* is defined by

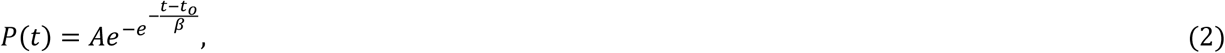

with initial condition *P(t_o_*) = *P_o_*, *A* is the carrying capacity, the maximum capacity of the environment here, *β* >0. Here, Eq. (2) divided by *L* corresponds to the CDF of the Gumbel distribution at point *t*. The PDF is simply obtained by differentiating the latter with respect to *t*.

Overall, the same process as the logistic function was performed with the Gumbel distribution’s PDF and CDF. The results indicate that using Gumbel is strongly preferred over the logistic, regardless of whether the Gumbel PDF (daily) or CDF (cumulative) is used. The parameter estimates for the total number of cases are no longer caught up within a week and even visually, the trajectory of the graph suggests paths for each country that are smoother and more accommodating towards future outcomes.

Regarding the parameter estimates, while the roles of “*A*” and “*t_o_*” are analogous to those of “*L*” and “*t*_o_” from the logistic function, respectively, the parameter “beta” plays a somewhat different role-as a slope/duration dual-function parameter which shrinks or stretches the curve. The Gumbel model incorporates some level of skewness which allows it to pick up broader variation in the data. Note that the standard errors are larger for the PDF based estimates, which is to be expected since it uses the volatile daily data as opposed to rather-stable cumulative data.(See Table 5, Table 6 and Fig.7.)

**Fig 7.**
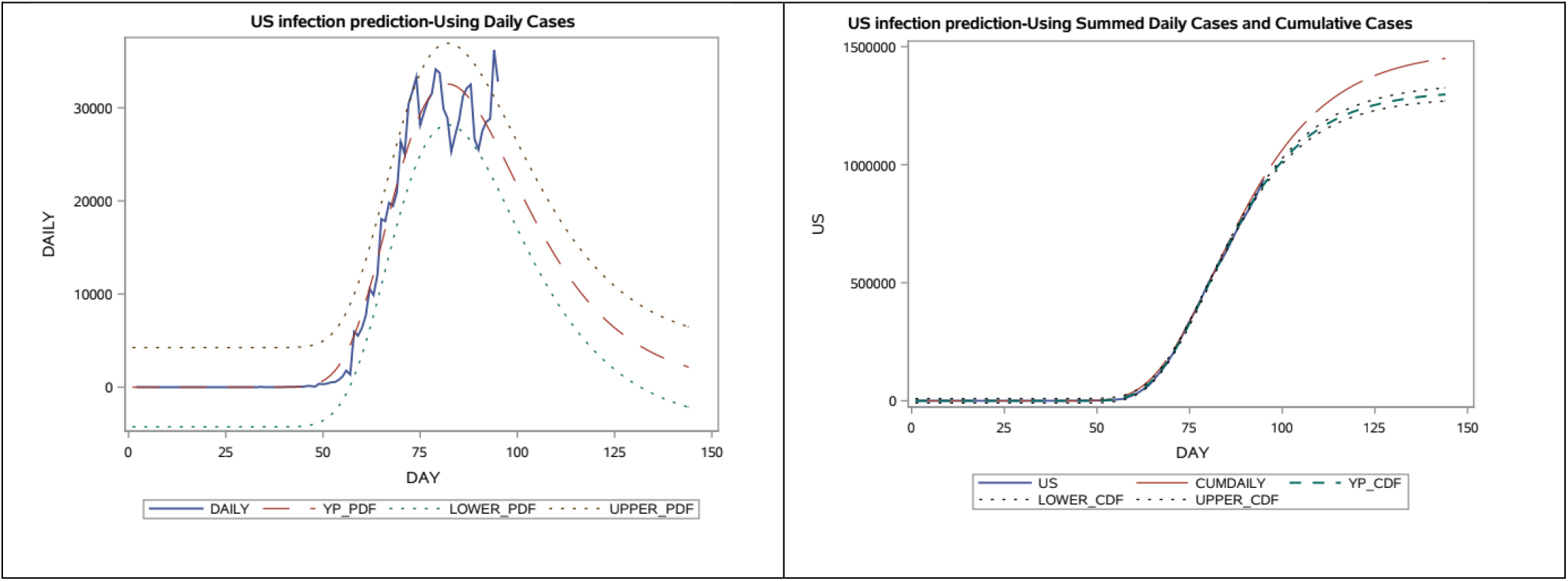
Observed daily US cases from 2020-01-22 to 04-25 and forecast for 7 weeks, using Gumbel PDF.

**Table 5.**
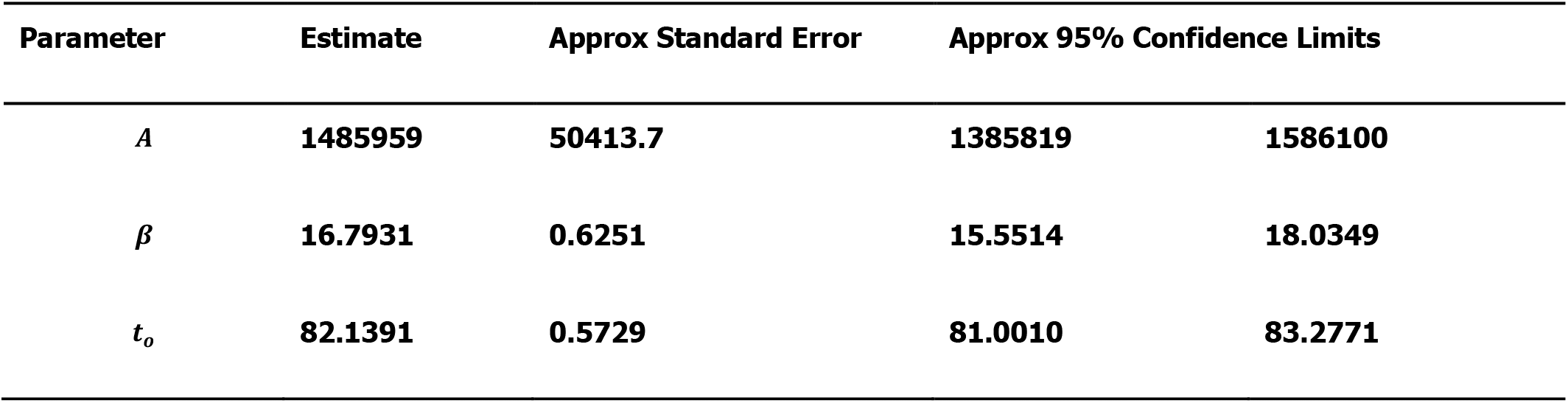
US- Gumbel PDF based parameter estimates.

**Table 6.** US- Gumbel CDF based parameter estimates.

| Parameter | Estimate | Approx Standard Error | Approx 95% Confidence Limits |  |
| --- | --- | --- | --- | --- |
| $L$ | 1315121 | 14613.8 | 1286097 | 1344146 |
| $\alpha$ | 14.8945 | 0.1874 | 14.5224 | 15.2666 |
| $t_o$ | 79.8614 | 0.1936 | 79.4769 | 80.2458 |

### 2.3 Comparison and discussion

The following plots (Fig 8.) summarise the key difference in using Gumbel distribution over Logistic distribution for the modelling of COVID-19 infection cases. The data used here is the number of cases in the US until 2020-04-25, where the circles represent the number of cumulative cases. The left panel shows the different models based on the Logistic function and the right panel shows the different models based on the Gumbel distribution’s CDF. The different lines indicate how many weeks’ worth of observations have been left out to simulate the results that were obtained in the past. On the left panel, it is clear that the Logistic model fails to capture an important trait in the data, hence it fails to keep up with the data. This is, as argued above, due to the asymmetric nature of the data. On the right panel, however, the Gumbel model is much more robust in picking up such trends. Though the prediction from 3 weeks ago has overestimated the number of cases, thereafter the estimates have remained rather stable and appear to be converging for the past 2 weeks.

**Fig 8.**
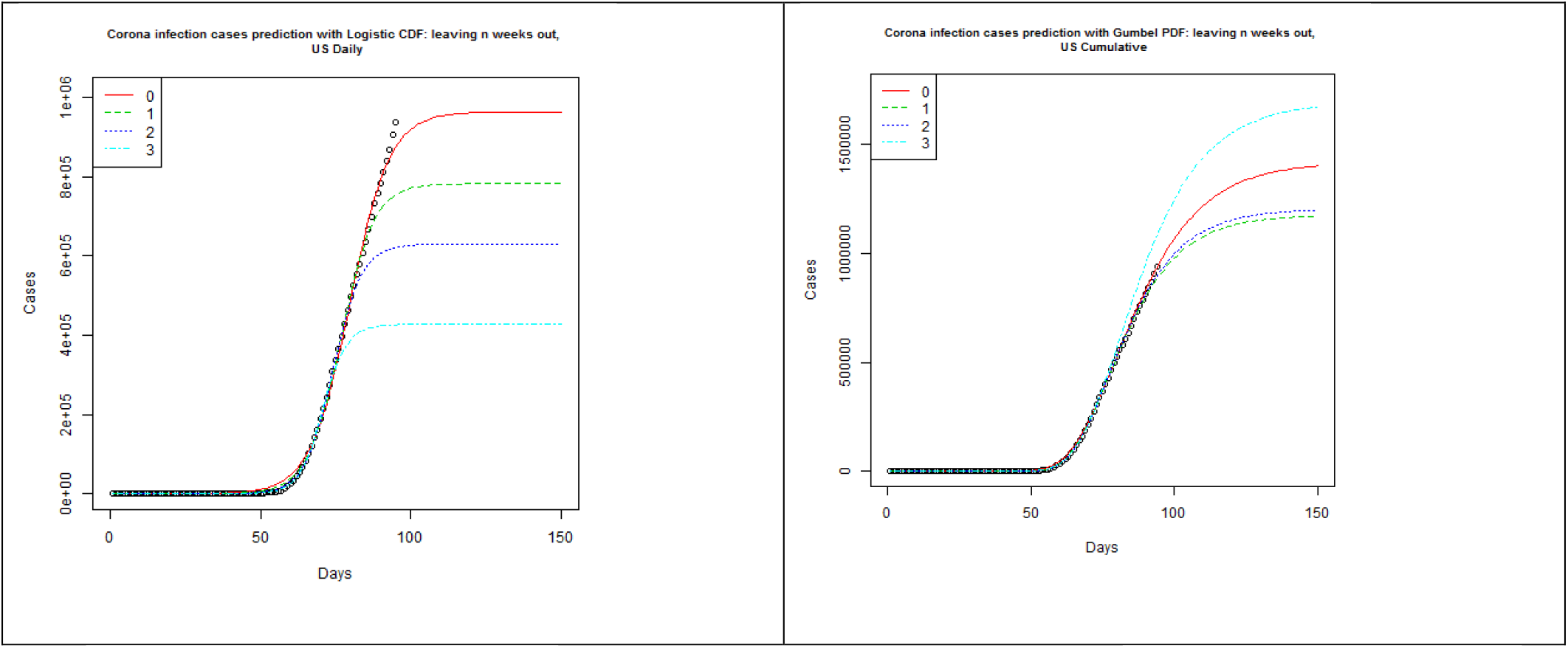
US- Logistic function and Gumbel PDF based forecasts from 2020-01-22 to 04-04 (3), 04-11 (2), 04-18 (1) and 04-25 (0)

## 3. Gumbel modelling for some selected countries

In this section, we analyse the dynamics of the coronavirus disease COVID-19 for some selected countries to show the potential of the Gumbel model (see Fig.9.). The time frame window is from 2020-01-22 to 04-04, 04-11, 04-18 and 04-25.

**Fig 9.**
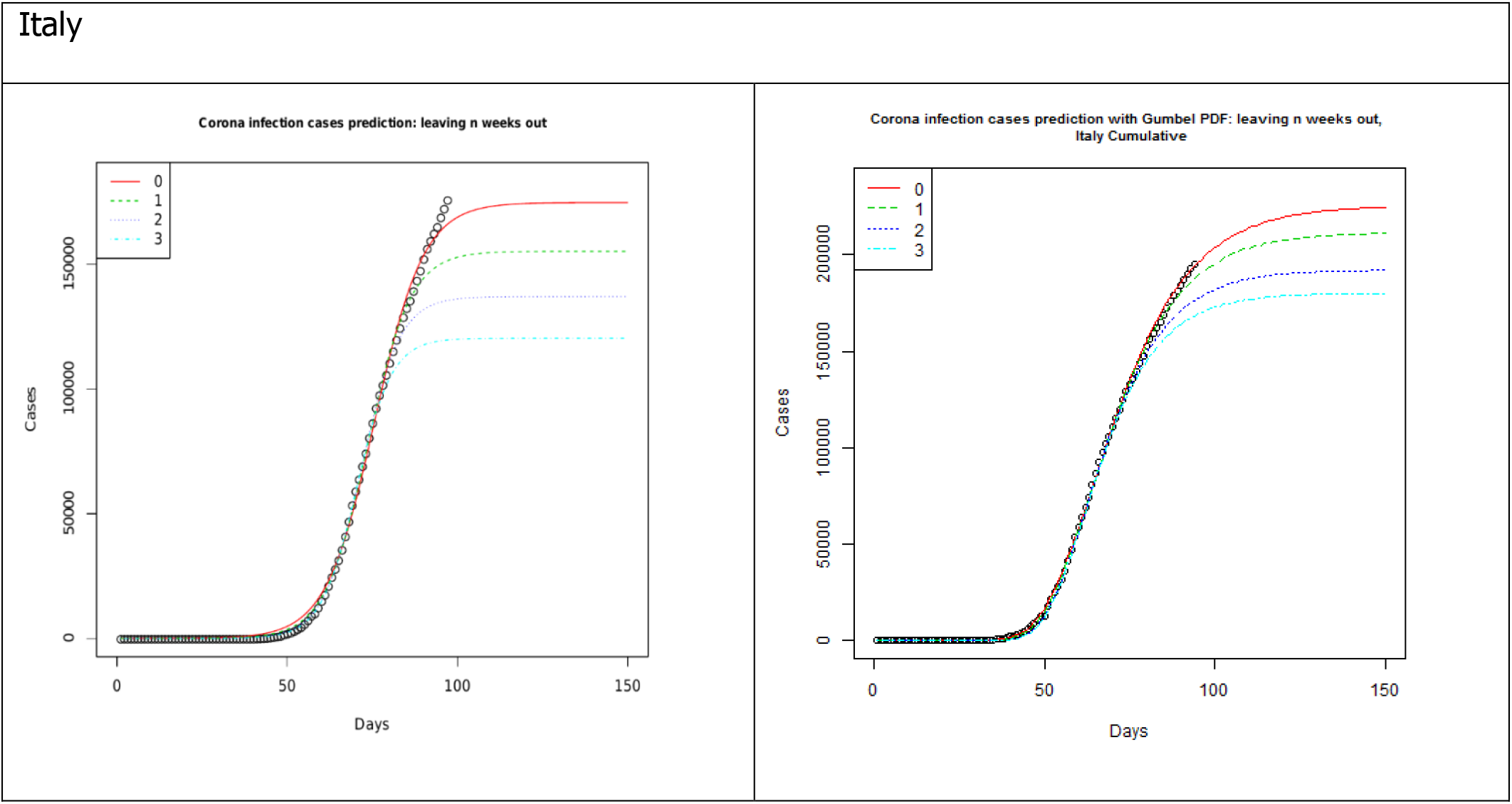
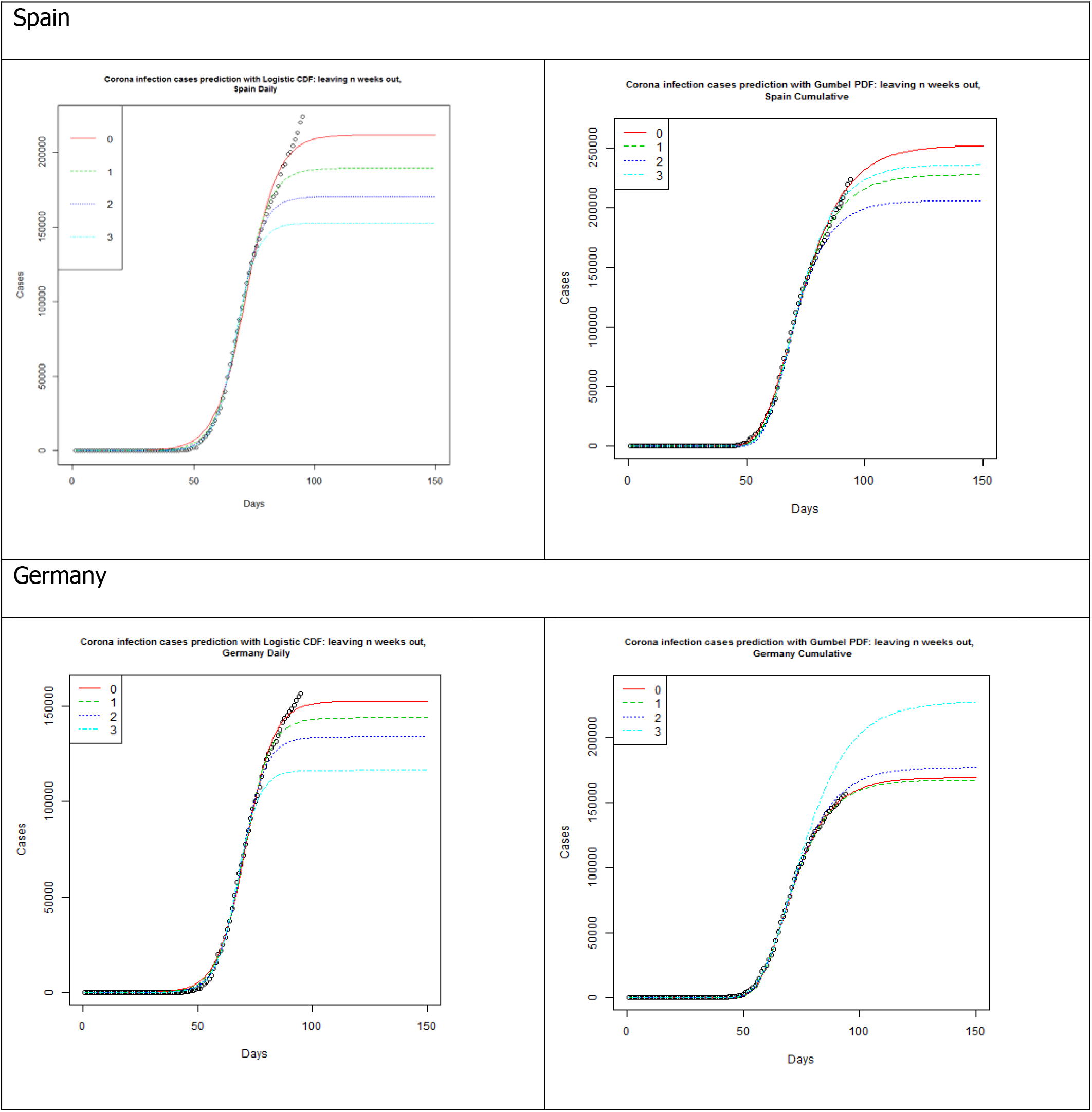
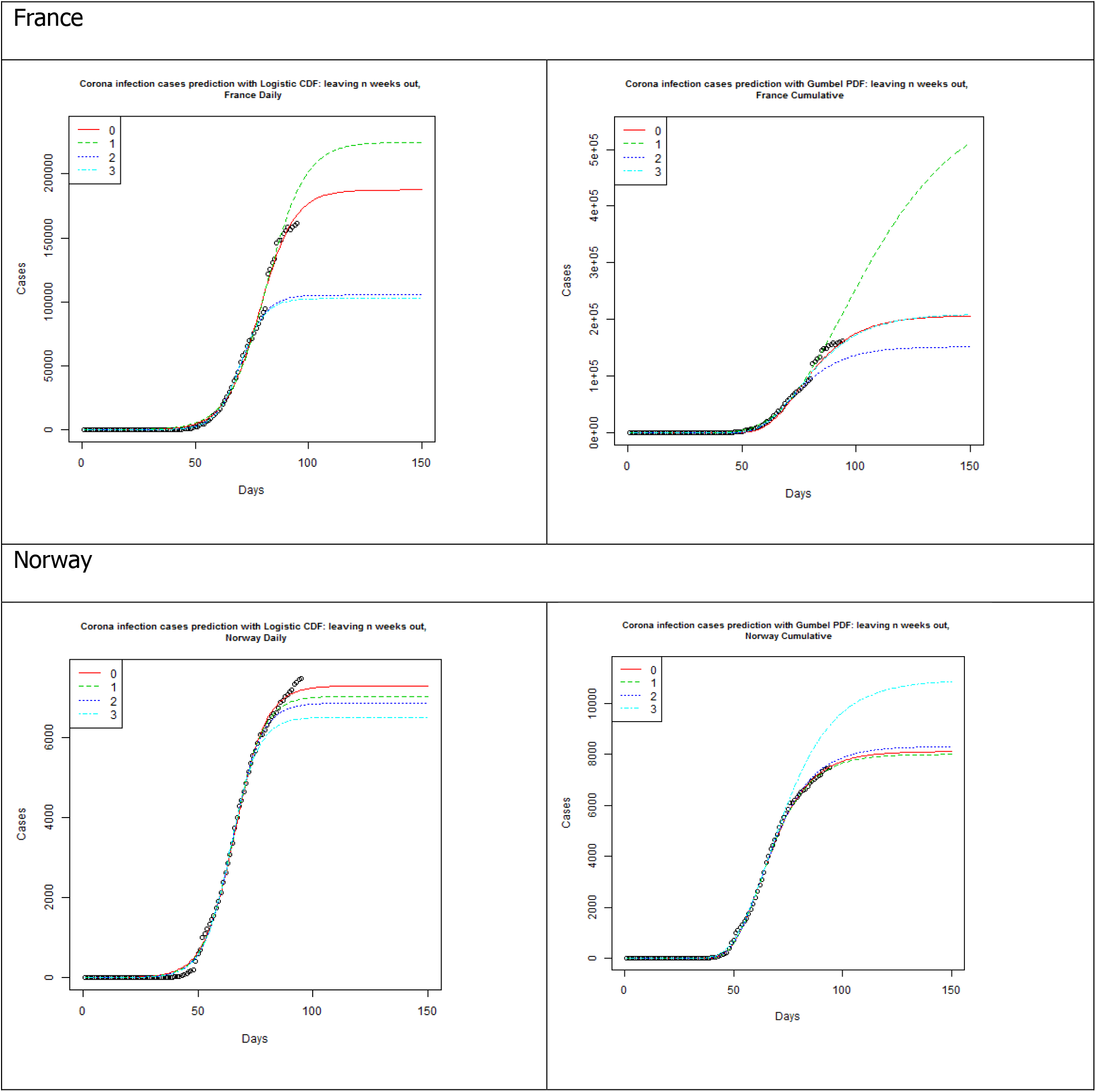
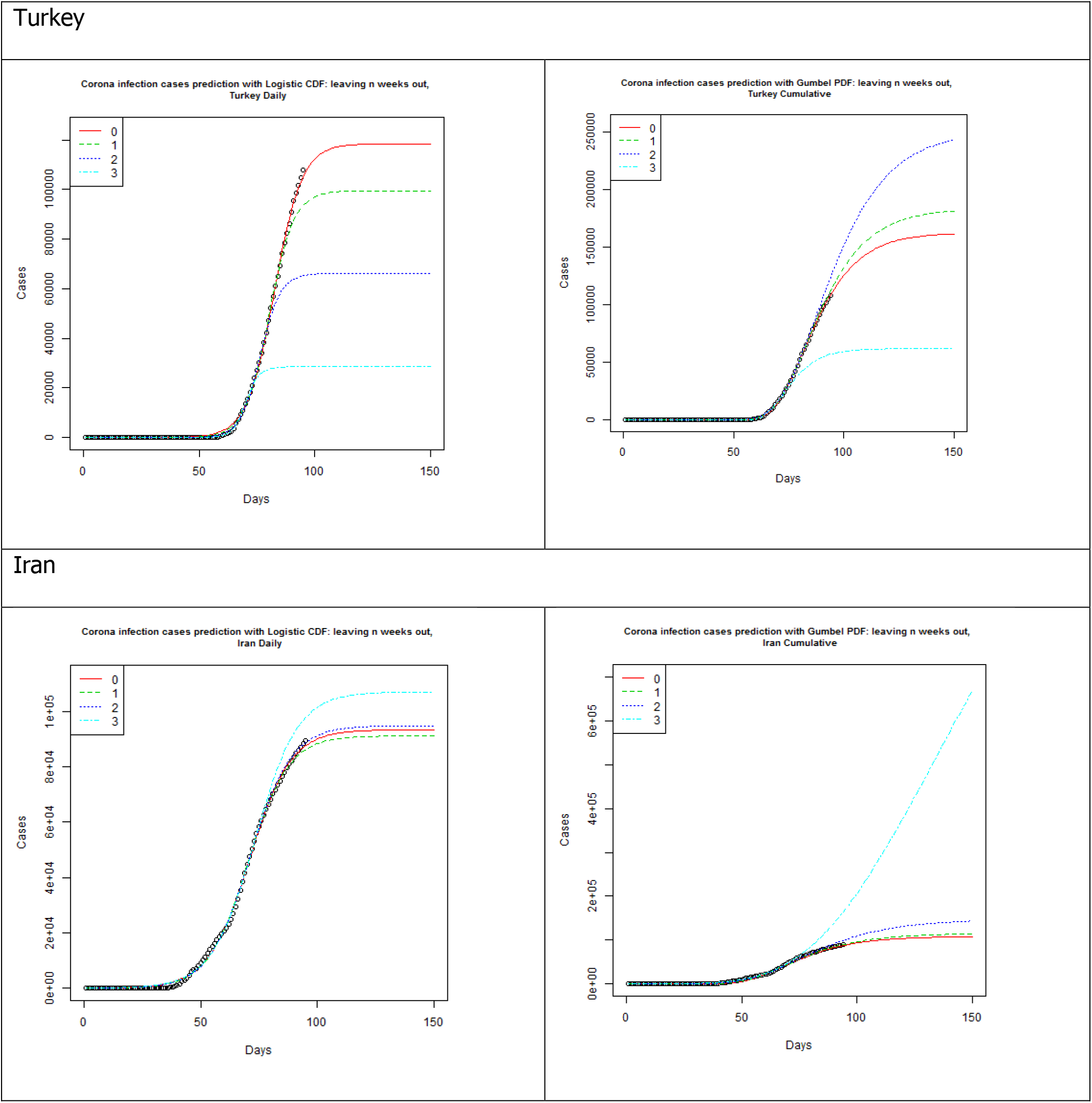
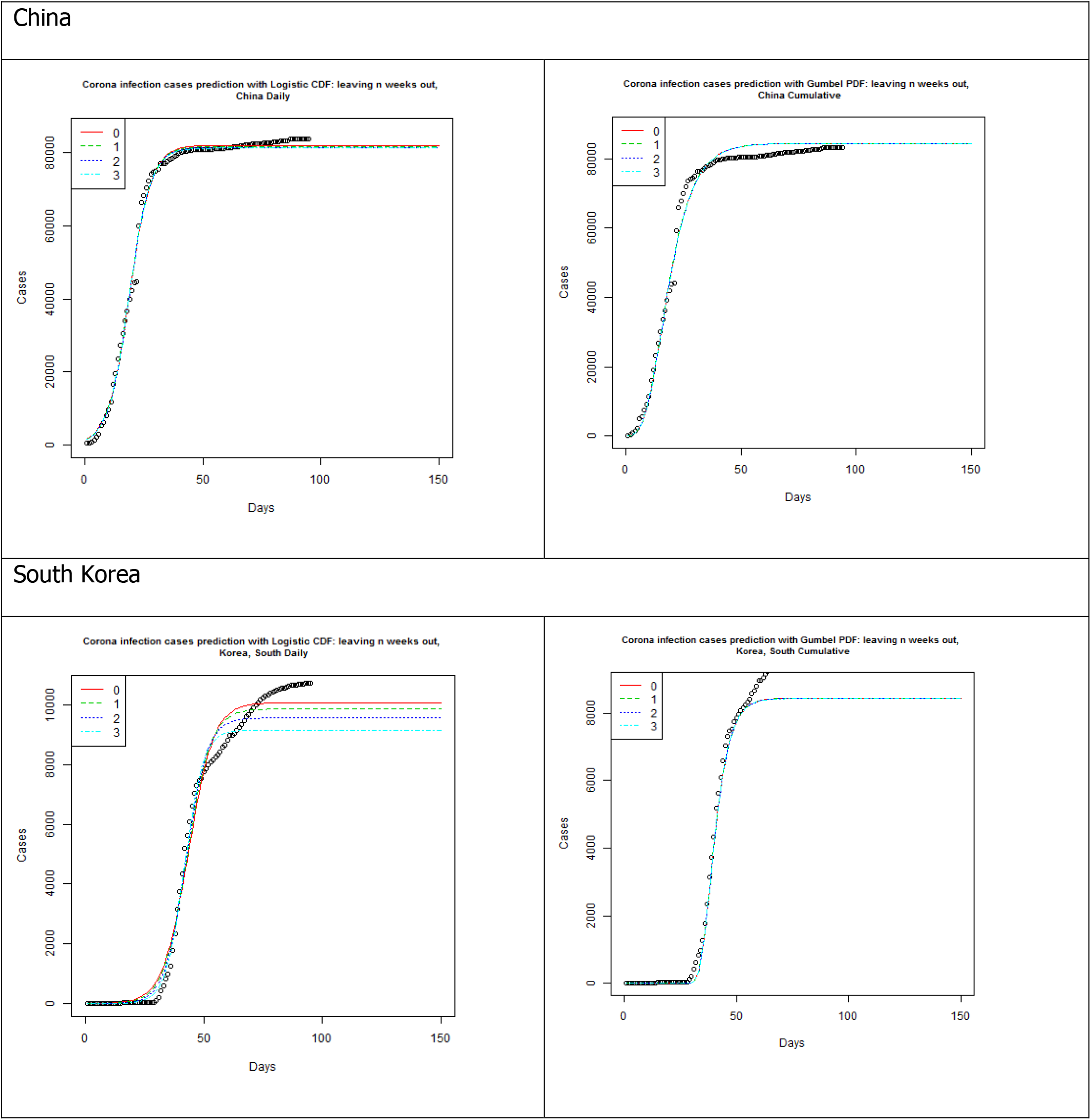
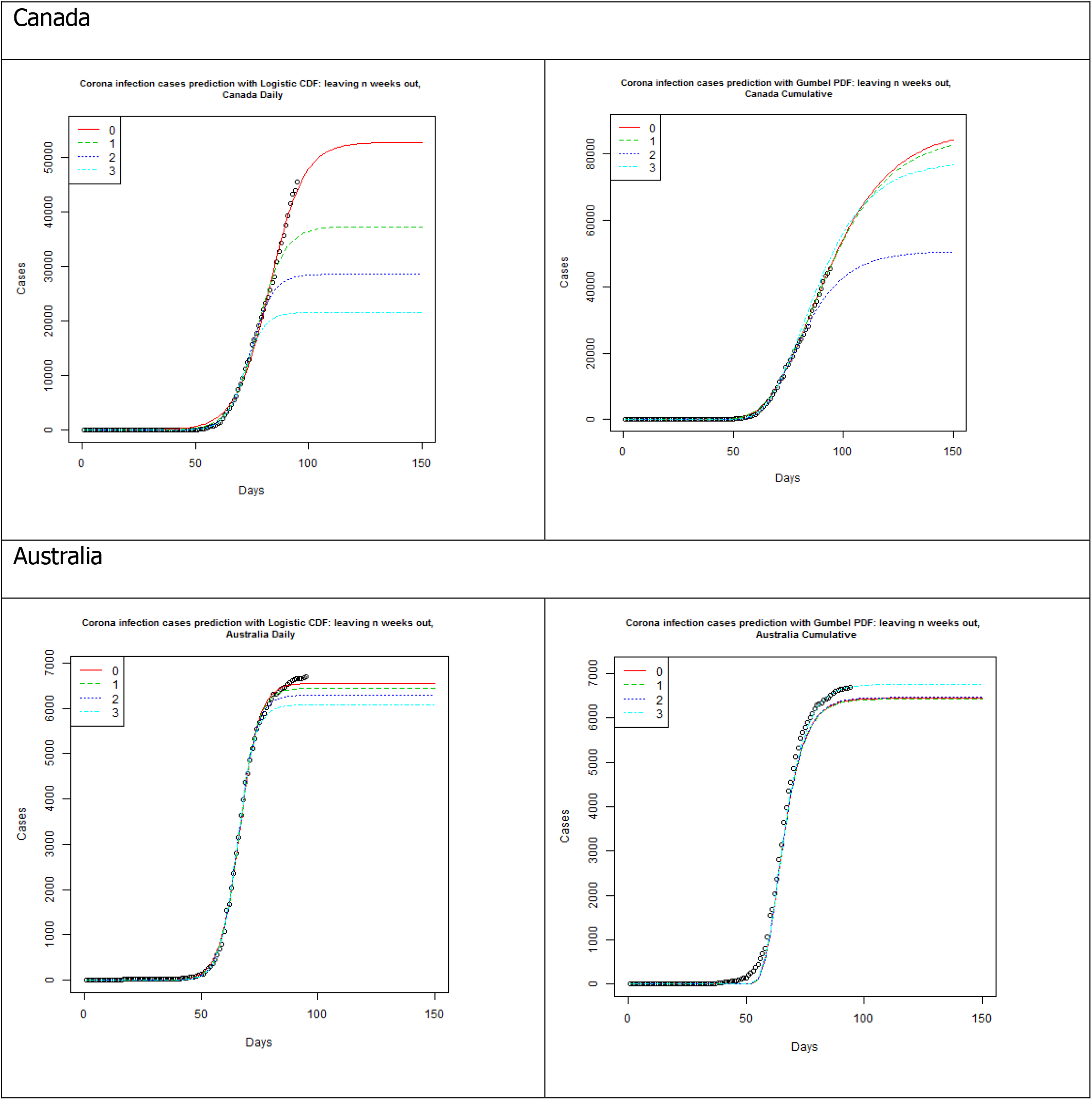
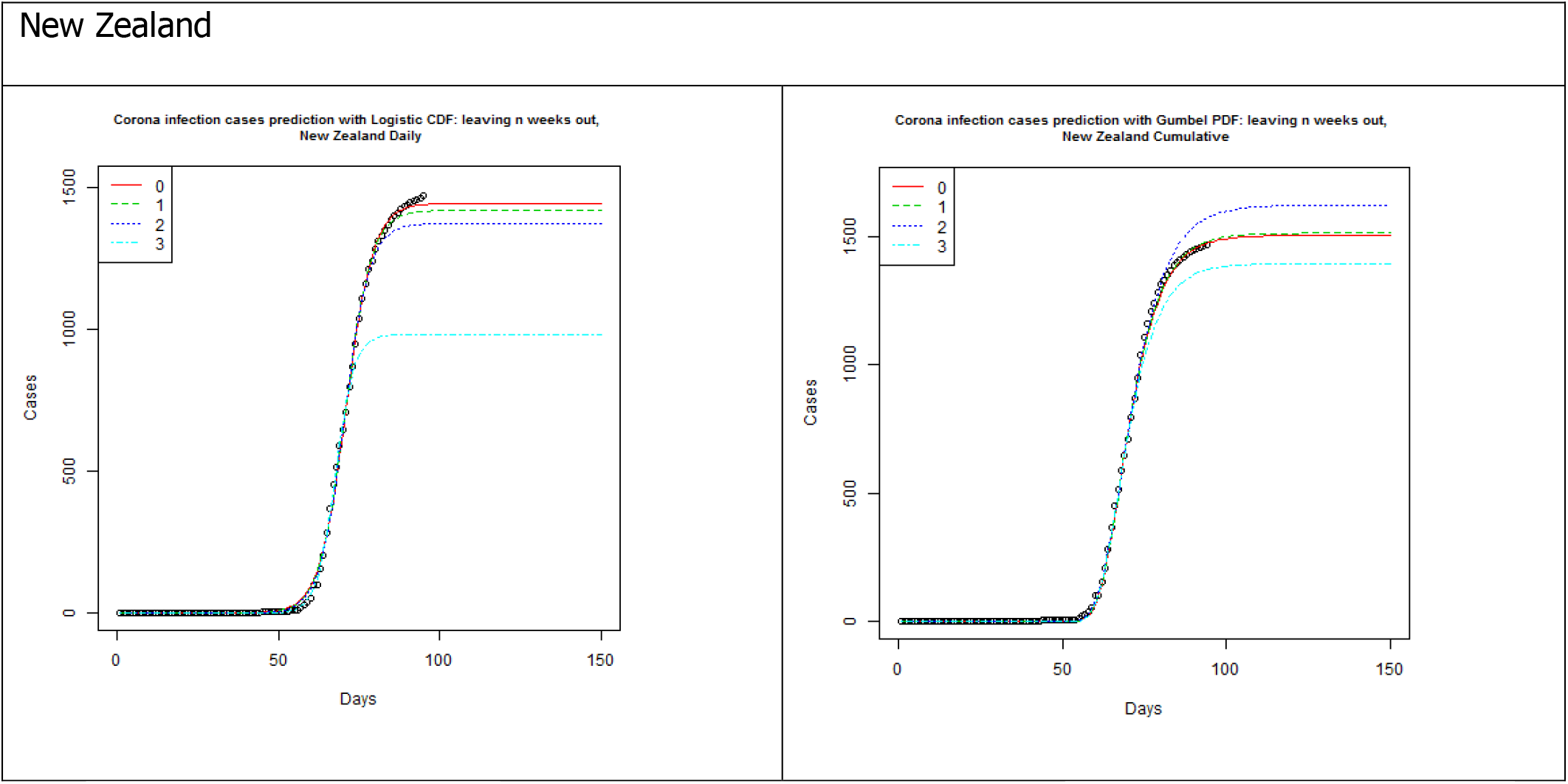
Logistic function and Gumbel PDF based forecasts from 2020-01-22 to 04-04 (3), 04-11 (2), 04-18 (1) and 04-25 (0), for selected countries.

## 4. Conclusions

In this paper, we have investigated the logistic growth model. The shortcomings were shown. We guided the reader to the solution of the use of the Gumbel model as an appropriate choice and completed the prediction for several countries. As [22] pointed out one model cannot answer all the questions. We hope this contribution can be a part of the set of solutions. The authors hope that this model will be of assistance for decision makers. This paper is part of an ongoing project related to modeling and prediction of the COVID-19 spread.

## Data Availability

There are a number of sources on the web that provide data on COVID-19 cases. One such site is The Humanitarian Data Exchange and one can find daily cumulative cases of COVID-19 per country. https://data.humdata.org/dataset/novel-coronavirus-2019-ncov-cases has a downloadable time_series_covid19_confirmed_global.csv starting from 2020-01-22, and for some countries, it even has the data broken down into different states or provinces. In order to perform the desired analysis, daily cases for each country had to be obtained, but some countries, such as the US and Australia, had the data broken down to state or provincial level. Since the focus of this research was per country, R open source software was used to sum along the unique values of Country, appropriately transforming the data for our analysis, then and non-linear regression was performed using the nls function.

## CRediT authorship contribution statement

Conceptualization: K.Y., M.A., and A.B.

Formal analysis: K.Y.

Funding acquisition: M.A. and A.B.

Investigation: K.Y., M.A., and A.B Methodology: K.Y.

Software: K.Y.

Supervision: M.A. and A.B.

Validation: M.A. and A.B.

Writing-original draft: K.Y.

Writing-review & editing: M.A. and A.B

## References

1. Little, N. COVID19Tracker.ca. 2020.

2. Chowell, G. Fitting dynamic models to epidemic outbreaks with quantified uncertainty: A primer for parameter uncertainty, identifiability, and forecasts. Infect Dis Model. 2017; 2(3), 379–398.

3. Viboud, C., Simonsen, L., Chowell, G.A. Generalized-growth model to characterize the early ascending phase of infectious disease outbreaks. Epidemics. 2016; 15, 27–37.

4. Chowell, G., Hincapie-Palacio, D., Ospina, J., Pell, B., Tariq, A., Dahal, S. Using Phenomenological Models to Characterize Transmissibility and Forecast Patterns and Final Burden of Zika Epidemics. PLoS Curr. 2016; 8.

5. Chowell, G. Tariq A, Hyman JM. A novel sub-epidemic modeling framework for short-term forecasting epidemic waves. BMC Med. 2019; 17(1), 1–18.

6. Chowell, G., Luo, R., Sun, K., Roosa, K., Tariq, A., Viboud, C. Real-time forecasting of epidemic trajectories using computational dynamic ensembles. Epidemics. 2020; 30, 100379.

7. Cohen, J. Scientists are racing to model the next moves of a coronavirus that’s still hard to predict. Science. 2020; DOI:10.1126/science.abb2161.

8. Batista, M. Estimation of the final size of the COVID-19 epidemic. medRxiv, 2020; 2020.2002.2016.20023606.

9. Roser, M., Ritchie, H., Ortiz-Ospina, E., Hasell, J. Coronavirus Pandemic (COVID-19). OurWorldInData.org. 2020; https://ourworldindata.org/coronavirus.

10. Hsu, J. Here’s how computer models simulate the future spread of new coronavirus. Sci. Am. 2020.

11. Anastasopoulou, C., Russo, L., Tsakris, A., Siettos, C. Data-based analysis, modelling and forecasting of the COVID-19 outbreak. PLOS ONE. 2020; 0230405.

12. Maier, B.F., Brockmann, D. Effective containment explains subexponential growth in recent confirmed COVID-19 cases in China. Science. 2020; eabb4557.

13. Cassaro, F.A.M., Pires, L.F. Can we predict the occurrence of COVID-19 cases? Considerations using a simple model of growth. Sci. Total Env. 2020;728, 138834.

14. Ceylan, Z. Estimation of COVID-19 prevalence in Italy, Spain, and France, Sci. Total Env. 2020; 729, 138817.

15. Salehi, M., Arashi, M., Bekker, A., Johan Ferreira, J.T., Chen, D., Esmaeili, F., Frances, M. A synergetic R Shiny portal to track COVID-19 demographic information, Submitted to Data Science Journal. 2020.

16. Sauer, N. Logistic growth and immunity. 2020; https://www.up.ac.za/media/shared/259/Documents/Covid-19/logistic-growth-and-immunity.zp189410.pdf.

17. Petropoulos, F., Makridakis, S. Forecasting the novel coronavirus COVID-19. PLOS ONE. 0231236 (2020).

18. Anderson, K.V., Daniewicz, S.R. Statistical analysis of the influence of defects on fatigue life using a Gumbel distribution. Int. J. Fatigue. 2018; 112, 78–83.

19. Gomez, Y.M., Bolfarine, H., Gomez, H.W. Gumbel distribution with heavy tails and applications to Environmental data. Math. Comp. Sim. 2019; 157, 115–129.

20. Hyun, N., Couper, D.J., Zeng, D. Gumbel regression models for a monotone increasing continuous biomarker subject to measurement error. J. Stat. Plann. Inf. 2019; 203, 160–168.

21. Huang, P., Hu, F., Dong, F. Parameter estimation of Gumbel distribution and its application to pitting corrosion depth of concrete girder bridges. Cluster Comp. 2019;22, S3405-S3411.

22. Panovska-Griffiths, J. Can mathematical modelling solve the current Covid-19 crisis? BMC Public Health, 2020; 20:551. https://doi.org/10.1186/s12889-020-08671-z.

